# Peripheral nerve conduction speed shows a disease control-dependent and -independent drop in type 1 diabetes mellitus in children

**DOI:** 10.1101/2022.12.08.22283120

**Authors:** Sarah S. Oberhauser, Dagmar l’Allemand, Jürg Lütschg, Philip J. Broser

## Abstract

**Background/Aim:** Nerve conduction speed (NCS) abnormalities are considered to be early signs of diabetic peripheral neuropathy. We investigated which determinants impact the NCS and how it is related to markers of metabolic control in children and young adults with diabetes mellitus.

**Method:** Fifty-four children aged five to 23 years suffering from type I diabetes mellitus were recruited into this study, which was conducted at the Children’s Hospital of Eastern Switzerland in St Gallen from March 2016 to June 2022. The metabolic control parameters were recorded and a nerve conduction study analyzing three motor nerves and one sensory nerve was performed. The data were compared to a control population of healthy children of the same height, and the height-adjusted NCS (dNCS) was analysed.

**Results:** For all four nerves under investigation, a statistically significant drop in the NCS of approximately 5 m/s independent of metabolic control was found, the peroneal nerve being the most sensitive. The NCS of the peroneal nerve correlated significantly negatively with the long-term haemoglobin with bound glucose (HbA1c) and highly significantly negatively with the standard deviation of mean glucose (SD), but there was only a trend with the HbA1c and the time in range (TIR) at the time of neurography.

**Interpretation:** All patients with diabetes mellitus showed a reduced NCS, partly independent of metabolic control. This may be due to a lack of the C-peptide, which regulates critical axonal membrane enzymes. High glucose variability clearly increases the risk of neuropathy, together with but also independently of the mean plasma glucose level.

## Introduction

Fast information transmission for perception and movement control in the human body is mediated by the peripheral nervous system. The information is sent via trains of action potentials propagating along the axons. In myelinated axons that are conducting the commands for muscle control and epicritical sensory perception, the action potential moves in saltatory fashion from ranvier node to ranvier node, achieving propagating speeds above 50 m/s. However, this form of long-distance communication is critically dependent on an individual’s well-controlled metabolic status. Small metabolic perturbations such as changes in electrolyte (Tinawi 2021), glucose levels (Klimontov 2021) or temperature (Denys 1991) lead to an immediate reduction in the nerve conduction speed (NCS). In addition to the acute effect, metabolic perturbations such as changes in the hormone level (i.e. insulin, C-peptide and fT4) or glucose levels can lead to functional and structural changes at the axons (Kamiya 2009;2009; Gupta 2016) or the myelin sheath, leading to a reduction in NCS. Thus, nerve conduction studies are a gold standard method for assessing peripheral neuropathy, which always begins subclinical (Hyllienmark 2013), progressing to clinically evident neuropathy with a cascade of increasing symptoms, decreasing the quality of life (Benbow 1998).

The pathogenesis of diabetic neuropathy has not yet been completely elucidated. Hyperglycaemia is the initiating factor of multiple pathways leading to osmotic and oxidative stress as well as inflammation, followed by dysfunction and structural change in the vascular endothelium and the Na,K-ATPase, an intermembrane pump critically important for nerve signaling, nerve conduction and kidney function (Vague 2004; Kallinikou 2019; Washburn 2021). The **polyol pathway** describes the conversion of glucose by aldose reductase to sorbitol and the oxidation to fructose. Its accumulation in the nerve surrounding tissues leads to low myoinositol levels and increased protein kinase C in nerves, causing a reduction in Na,K-ATPase function (Yagihashi 2010). Glycosylated myelin is endocytosed by macrophages (Vlassara 1985). Further, chronic hyperglycaemia induces the formation of AGEs (advanced glycation end-products), which can modify myelin as well as cytoskeletal proteins, such as tubulin, actin and neurofilaments, leading to axonal degeneration and dysfunction (Verotti 2001). Once the AGEs bind at their receptor (RAGE), they promote transcription of proinflammatory genes, leading to oxidative stress (Bierhaus 2001, 2004). The **metabolic flux hypothesis** highlights the pathogenic potency of the cofactor turnover of NADPH to NADP+ and NAD+ to NADH in the polyol pathway. This cofactor turnover is strongly associated with the development of oxidative stress (ROS = reactive oxygen species), leading to cell dysfunction, reduced energy level and cell apoptosis. Furthermore, the **hypoxia hypothesis** describes low nitric oxide (NO) levels with vasoconstriction and low perfusion of nerves and other tissues due to hyperglycaemia-induced inhibition of the endothelial nitric oxide synthase (eNOs). This imbalance is reinforced by overproduction of vasoconstrictor substances such as endothelin-1 in hyperglycaemia. (Kallinikou 2019)

**C-peptide**, a cleaved portion of the insulin prohormone, was long thought not to have physiological effects. Nowadays we know that it stimulates the Na,K-ATPase and the endothelial nitric oxide synthase (eNOS), both deficient in diabetes disease. Beneficial effects are explained partly by better microcirculation, but also by a partial reverse of acute and chronic metabolic, functional and structural changes in peripheral nerves, as particularly demonstrated in laboratory and clinical studies (Vague 2004; Kamiya 2009 Fall; Waschburn 2021). The latter have shown an amelioration of the autonomic and sensory neuropathy as well as nephropathy (Ekberg 2003; Johansson 2000). However, the fact that Ekberg et al. did not find an amelioration of the peroneal NCS after three months of C-peptide treatment may be explained by different mechanisms underlying the conduction defect in sensory and motor nerves, the higher sensitivity of the peroneal nerve function being altered by hyperglycaemia, the rather short treatment time, treatment for neuropathy pathogenesis that was started too late and insufficient glycaemic control screening using only HbA1c measurement.

Genetic variant polymorphisms of the aldose reductase and oxidant enzymes (Kallinikou 2019) as well as the individual duration of the remission phase with secretion of the protective C-peptide may explain an individual susceptibility for developing a neuropathy.

In patients with diabetes mellitus (DM), both types of interferences, functional and structural, are present. Glycaemic control through insulin replacement can reduce diabetes-associated vascular complications, but even intensive glycaemic control does not normalize the risk of developing comorbidities (Nathan 1993). The plasma glucose levels still show much more variation in comparison to healthy individuals, and the hormone status is altered with an external supplementation of insulin and the absence of C-peptide, which has a critical effect on the Na,K-ATPase (Johansson 2000; Vague 2004; Washburn 2021).

Especially during the first years of the disease, it is expected that the effect on the nervous system is primary related to the direct metabolic effects and much less related to long-standing complications such as structural changes leading to micro- and later macroangiopathy.

This study was performed in order to understand the effects of T1DM on the peripheral nervous system during the first years after disease onset in children and young adults.

## Method

This prospective, cross-sectional study was conducted at the Children’s Hospital of Eastern Switzerland in St Gallen from March 2016 to June 2022. The study was approved by the ethics committee of Eastern Switzerland (Ethikkommission Ostschweiz, approval no: 22/018) and registered at the Swiss project database (2022-00216). Written informed consent was obtained from the caregivers before the inclusion of participants.

Since measurement of NCS is possible from preschool age on, children and young adults diagnosed with T1DM at or after the age of five years were eligible for inclusion into the study. To avoid disturbances of the disease manifestation with osmotic, fluid and electrolyte imbalances, the minimum time from DM manifestation had to be six months. Children with other chronic diseases, premature birth, severe acute disease or a family history of any inherited neurological disease were excluded. Children were randomly recruited at the outpatient diabetes clinics.

### The ‘St Gallen diabetes programme’

According to the ‘Sankt Gall diabetes programme’ guidelines, all patients with T1DM and MODY were seen three-monthly by a paediatric diabetologist for a short check-up and once a year for a general check-up for complications and comorbidities (Pihoker 2018), including a whole paediatric and neurological examination in which the vibration threshold was tested. Each visit included measurement of height, weight, blood pressure and HbA1c as well as an analysis of the glucose profile, with the characteristics time in range (TIR), mean glucose, incidences of ketoacidosis, hyperglycaemia (>10 mmol/l), hypoglycaemia °II (<3 mmol/l) and glucose variability (coefficient of variation, standard deviation of mean glucose). The glucose profile calculations for glucose measurements via sensor or blood glucose meter were done by the evaluation programs Diabass, Medtronic, FreeStyle Libre or Dexcom. For all patients the diabetes auto-antibodies (GAD, IA2, ZnT8, ICA and/ or IA) were measured to classify the subtype of diabetes. In the absence of diabetes-specific antibodies, MODY genetic was performed. In addition, a screening for autoimmune thyroid disease (AITD) and coeliac disease (CD) was done yearly.

Every second year, the annual check-up included a nerve conduction and a nerve ultrasound study in the neurophysiology department of the Children’s Hospital of Eastern Switzerland. This examination follows a strict protocol. First, the patients adapts to the ambient room temperature in the waiting area. In the waiting area the temperature is kept constant throughout the year. Then, a motor nerve conduction study of the peroneal nerve, the tibial nerve and the median nerve and a sensory nerve conduction study of the median nerve are conducted serially, according to the clinical standards of Broser, Lütschg 2020. In all patients, the right side is tested, except where this is not possible (e.g. injury to the right side). In addition, high-resolution, cross-sectional ultrasound (Canon Aplio i800, Japan) images of the left median nerve at the level of the wrist are taken (data not part of this study).

### Data collection

In order to correlate the diabetes data with the nerve conduction data, the following procedure was performed. A period of 90 days before the NCS study – coinciding with the time period of the HbA1c – was selected as the relevant period. The data stored in the continuous glucose monitoring (CGM) program or the program of the blood glucose meter with the abovementioned variables were analysed. In addition, correlations were sought between the NCS and autoimmune status as well as the age, sex, duration of diabetes disease, weight, height and comorbidities. Long-term metabolic control was assessed by the mean HbA1c, calculated from the HbA1c values of the last five annual check-ups, and the incidence of ketoacidosis and hypoglycaemia °III since disease onset.

### Control group

The department of paediatric clinical neurophysiology at the Children’s Hospital of Eastern Switzerland continuously collects normative control data. The normative controls are recruited from a large group of children with mild, unilateral, traumatic nerve injuries. These otherwise completely healthy children and their caregivers were asked to volunteer. If consent was given, the same measurement protocol as described above was performed. However, the unaffected body side was chosen (for details see Broser 2020). The complete list of data of the control group is given in Supplementary Table 3.

### Height adjustment for the peroneal nerve

It is well documented in the literature that the NCS of the peroneal nerve is negatively correlated with body height, registering a decrease of around 0.065 m/s/cm (Hyllienmark, Ludvigsson, Brismar 1995). Therefore, we included data from a control group to adjust the NCS for height, using the formula: difference (measured NCS – NCS of healthy children of the same height) = **dNCS**.

### Processing and statistical computation

An a priori power calculation estimated a minimum number of 50 diabetic patients and 20 control patients to achieve statistical significance. All patient data were entered into an Excel sheet for data storage. All statistical computation, processing and figure generations were performed in R (R statistical computation).

Two-sample Wilcoxon tests (Mann–Whitney Test) or Kendall Tau were used to perform group comparisons (Hollander & Wolfe 1973). To correlate the metabolic control parameter with the NCS, Pearson correlation tests were performed. The significance level, α, was set to *α* = 5%. In this study, a total of eight statistical tests were performed. In order to correct for multiple comparisons, Bonferroni’s method (Benjamini 1995) was applied.

## Results

### Subjects

Altogether, 58 patients were screened for inclusion into the study. Data were complete, and consent for inclusion was given by 54 patients. The gender ratio was 28 males (52%) to 26 females (48%) and did not influence the NCS. The study group had a median age of 14 years (range 6–23 years) and a median disease duration of six years (range 0.5–18 years). Of the patients, 89% had at least one positive diabetes-specific antibody, whereas in 7% MODY was diagnosed. Continuous glucose monitoring (CGM) was used in 70%, an insulin pump in 52%. The median HbA1c was 8% (range 5.6–12.9%). A summary of the patients’ characteristics is given in Table 1, a complete list of the subjects is presented in Supplementary Table 1.

**Table 1.** Summary of study data

### Diabetes group vs. control group

The NCS of the three motor nerves and one sensory nerve were compared for the study and control group (see Figure 1, Supplementary Table 2). All four tests showed statistical significant differences, the NCS of the diabetic group being about 3–5 m/s slower than that of the control group. The peroneal nerve was the most sensitive nerve (see Supplementary Figure 1), showing a highly significant reduction compared to the control group (p value < 0.0001). On the basis of the generally used peroneal NCS limit of 50 m/s, 75% of our study group patients showed a pathology - even with a median age of only 14 years and a median disease duration of only six years. The raw NCS data of the peroneal nerve showed a decline in the NCS with age for both groups (Supplementary Figure 1). Further analysis attributed this decline of NCS to the increase in body height (see Supplementary Figure 2). In order to take this physiological drop in NCS into account, the difference between the measured and the expected NCS was calculated (dNCS) and used for further analysis. As a result, the significant correlation between age (p = 0.002) and the NCS no longer persisted. The heights of the males and females in our study group were not significantly different; nor were the evaluation of the NCS and the dNCS sex-specific.

**Figure 1:** Panels A to D, boxplots for the NCS of the control and diabetes groups. The analysed nerve is written above each panel. Stars indicate the significance level.

### Metabolic control and NCS

The data from the study suggest only a trend between the peroneal dNCS and the TIR in patients with a CGM wearing time of >/= 70% as well as the HbA1c (Figure 2, Panels A and B). However, the mean HbA1c of the last five years showed a significant negative correlation with the peroneal dNCS (R = -0.37, p = 0.011*) and the glucose variability expressed by the coefficient of variation was significantly negatively, the standard deviation of mean glucose highly significantly negatively correlated with the peroneal dNCS (Figure 2, Panels C and D). In contrast to the HbA1c and the TIR, the mean glucose also correlated significantly negatively with the dNCS (R = -0.33, p = 0.023).

**Figure 2:** Panels A to D, scatter plots relating a metabolic control parameter on the X axis to the residual of the NCS of the peroneal nerve. A Pearson correlation is calculated for each graph. The result is plotted in the upper left corner.

### Effect of disease onset and duration

The data of this study did not show an obvious correlation between a drop in the dNCS and the duration of diabetes or age of diabetes onset. In Figure 3, Panels A and C, the data points are coloured based on the glucose SD values; in Panels B and D, the points are coloured based on the mean HbA1c values for the last five years. Figure 3, Panels A and B, show the dNCS in relation to the age at onset of diabetes disease. In neither panel is a trend visible, suggesting that at least no obvious relation between the dNCS and age at onset exists, either before or after puberty. Figure 3, Panel C and D, show the dNCS in relation to the duration of diabetes disease. No obvious trend was found. However, during the first five years after diabetes onset, the peroneal dNCS decreases and reaches a steady state at about 5 to 10 m/s reduction (Supplementary Figure 4).

**Figure 3:** Panels A and B, scatter plots relating the age of onset on the X axis to the dNCS of the peroneal nerve. In panel A, the dots are coloured according to the standard deviation of mean glucose. In panel B, the dots are coloured according to the mean HbA1c. In panels C and D, the scatter plots relate the duration to the dNCS.

### Autoimmunity and reduction in NCS

T1DM is an autoimmune disorder and is associated with other autoimmune diseases such as coeliac disease (CD) and autoimmune thyroid disease (AITD). In this disorder, a variable number and quantity of diabetes-specific antibodies (GAD, IA2, ZnT8, ICA, IAA) are present. To explore if specific antibodies alter the effect of hyperglycaemia on the development of diabetic peripheral neuropathy (DPN), graphs relating to dNCS, diabetes control and antibody status were plotted (see Supplementary Figure 3). No obvious effect could be found, neither for one of the five diabetes-specific antibodies GAD, IA2, ZnT8, ICA or IAA nor for the associated autoimmune diseases CD or AITD.

## Discussion

The incidence of diabetes mellitus is increasing worldwide (Patterson 2018) (DIAMOND Project Group 2006). The type 1 diabetes mellitus (T1DM) goes along with the increasing number of autoimmune diseases, Type 2 diabetes mellitus is a complication of a rise in the incidence of obesity. Diabetes mellitus (DM) is associated with a significant overall disease burden, primary vascular complications and nephropathy, retinopathy and neuropathy as consequences of microvascular damage.

Symptoms of these complications are rarely seen in children and adolescents (Hyllienmark, Brismar, Ludvigson 1995), but there is evidence that their pathogenesis and early signs develop already during childhood (Marcovecchio 2011). In our cross-sectional study, we showed that the NCS is approximately 3–5 m/s lower in the diabetic group compared to the non-diabetic control group. This reduction was independent of glycemic control and was not associated with clinical symptoms such as lower limb pain, paresthesias, and/or hyperhidrosis; only in a few patients was a reduced vibration threshold noted.

The drop in NCS was similar for all three motor nerves and the sensory nerve, but the most consistent loss in NCS was found for the peroneal nerve (Figure 1). This is also similar to the findings of other studies (Riihimaa 2001) (Hyllienmark, Brismar, Ludvigsson 1995).

It is well documented in the literature that the NCS correlates negatively with body height in both diabetic and healthy persons (Louraki 2012; Hyllienmark, Ludvigsson, Brismar 1995). In order to normalize for body height, we calculated the loss of nerve conduction speed (dNCS) in relation to the healthy control group. We used our own control group to calculate the linear fit, which wavery close to the model presented by Hyllienmark, Ludvigson, Brismar (1995) (Supplementary Figure 2, black and blue line). This normalization has the advantage of equalizing for body height dependent differences in NCS. Using this height-adjusted evaluation of NCS (dNCS), the data showed that neither age at the time of disease onset nor the duration of the diabetes was correlated with a loss of NCS (Figure 3). The **age effect** at disease onset and in general on the degree of DPN was discussed controversially in the past. Some researchers observed that patients aged 5–14 years at the time of T1DM onset had an increased risk of complications compared with those diagnosed at either a much younger age or after puberty (Monti 2007). Our study group with a disease onset ranging from 1.5 to 15 years did not show this relationship, in agreement with the findings of Ludvigsson et al. (1979). **Sex as a risk factor for DPN** was also controversially discussed. Variably, females (Monti 2007) and males (Olsen 2000) are thought to be more vulnerable. Our results were sex-neutral, like other studies (Solders 1997). **Puberty** is thought to influence the course of diabetic microvascular complications. Marcovecchio et al. (2011) and Riihimaa et al. (2001) describe puberty as an adverse event, whereas others found late puberty to be a risk factor for DPN (Barkai 2000). Our study group, with an age range of 1.5 to 23 years, did not show any clear trend for NCS to be worse during or after puberty, which normally occurs between the ages of 10 and 16 years.

When Solders at al. (1997) conducted a 10-year follow-up study in newly diagnosed children with T1DM, they found a reduced nerve conduction velocity, which improved over the **first two years**. Their longitudinal study with neurophysiological recordings of nerve conduction after two, five and 10 years identified further deteriorations over time. Our study group showed a continuous significant reduction of dNCS during the first five years of disease, regardless of glycaemic control. We interpret this as a ceased diabetes remission, along with a gradual end of the secretion of C-peptide. In several animal experiments, the C-peptide showed an effect on the NCS; in addition, it is known from in vitro studies that the C-peptide has a regulatory effect on the activity of the Na,K-ATPase. Reduced activity may explain the loss in NCS (Vague 2004; Kamiya 2009 Fall; Waschburn 2021).

Our data suggest that, after five years of diabetes disease, the glycaemic control is the dominant factor for nerve function (Figure 2). This result is in line with the data by Ziegler et al. (2015) and the Diabetes Control and Complications Trial (Nathan 1993) and highlights the importance of **glycaemic control**. Ziegler et al. found a significant decrease in NCS with a higher mean HbA1c. Our data also show a negative correlation between mean (over five years) HbA1c and HbA1c at the time of neurography with a drop in dNCS. For example, an increased HbA1c goes along with a decreased dNCS of 3–5 m/s. However, the correlation between HbA1c and dNCS was rather weak and, even more surprisingly, the TIR in >/= 70% CGM-wearing patients (Figure 2) showed only a slight trend.

Given that the majority of patients in this study had a glucose sensor, it was possible to record and analyse the precise glucose profile for a period of 90 days before the nerve conduction study. This time period was chosen in accordance to the reflected time span of 2-3 months by the HbA1c. From these data, the mean glucose (mean), the standard deviation (SD) and the coefficient of variation (CV = (SD / mean glucose) * 100) were calculated. Astonishingly, and in contrast to the simple HbA1c and TIR, a significant correlation was found between dNCS and the mean glucose (p = 0.023) and the glucose variability, expressed by the coefficient of variation and the standard deviation of mean glucose (Figure 2). The correlation was very strong for the standard deviation of mean glucose, providing increasing evidence that glucose variability is an independent risk factor for DPN; the effect of the overall mean glucose level was, however, less (Klimontiv 2021).

The observation period of the study was too short to quantitatively determine the influence of the **treatment type** (multiple daily insulin injections, insulin pump, CGM use, closed loop). However, the data showed no difference in dNCS according to the type of treatment.

For completeness, we tested whether different autoantibodies had an effect on the dNCS. We did not find any differences.

We conclude that DM disease leads to a glycaemic control-dependent and -independent drop in NCS, as a first sign of DPN. To exclude height from the evaluation, height-adjusted NCS should be analysed. The pathogenesis of DPN is still not completely understood. Our study showed that microvascular complications such as neuropathy begin early and are dependent on long-term metabolic control according to individual susceptibility. Thus, it is recommended that diabetic patients are screened regularly, i.e. with yearly clinical examinations and a nerve conduction study every second years. At-risk patients should be identified early, and glycaemic control should be optimized, because DPN can be reversible at a young age (White 1981) or at least its progression slowed down (Nathan 1993; Ziegler 2015). Besides, in future, it is possible that therapeutic options such as C-peptide will be available; where they are available, should be given early in the course of DPN disease.

### Strengths and Limitations

This was the first study analyzing the height-adjusted reduction of NCS in diabetic patients to exclude height as a bias in the evaluation of influencing factors like age and duration of disease. A follow-up study with longitudinal NCS examination and parallel C-peptide measurement is planned.

Several data problems were present, such as inconsistently standardized analysis of the glucose profile; in particular, the sensor-wearing time should be known well when analyzing the TIR value. This value may possibly have a correlation with the NCS when evaluated over a long time such as five years, like the mean HbA1c.

The size of the study was too small to perform subgroup analysis and to test if further factors are relevant for the development of a diabetic neuropathy. The neurophysiological examination focused on the NCS, but other functions of the motor and sensor control, e.g. neuromuscular transmission, may also be affected.

## Data Availability

All data produced in the present study are available upon reasonable request to the authors.

## Abbreviations

AITD: autoimmune thyroid disease
CGM: continuous glucose monitoring
CMAP: compound muscle action potential
CD: coeliac disease
CV: coefficient of variation
CSII: continuous subcutaneous insulin infusion
DM: diabetes mellitus
DPN: diabetic peripheral neuropathy
dNCS: residual drop in the peroneal NCS after correction for body height
GAD: glutamic acid decarboxylase antibody
glucose SD: standard deviation of mean glucose
HbA1c: haemoglobin with bound glucose
IA: insulin antibody
IA2: tyrosine phosphatase 2 antibody
ICA: pancreatic cell antibody
MODY: maturity onset diabetes of the young
NCS: nerve conduction speed
R: degree of regression
SD: standard deviation
T1DM: type 1 diabetes mellitus
TIR: time in range
y: years
ZnT8: zinc transporter 8 antibody

## Acknowledgements

We thank: the FND team for their support in conducting the NCS measurements; the secretary of the KER team for organising the NLG clinic; and the Diabetes Team at the Children’s Hospital of Eastern Switzerland for their support. Finally, we wish to thank all the children who participated in the study.

## Conflicts of interest

The authors report no conflicts of interest.

## Figure and Table legends

Supplementary Figure 1:

Panels A to D, scatter plots relating the age to the NCS for each group, black representing the control group, green the diabetes group. The relevant nerve is named above each panel.

Supplementary Figure 2:

Scatter plot relating, on the X axis, the body length and, on the Y axis, the absolute NCS of the peroneal nerve. The black line is a linear regression line (p-Val = 0.013). The blue line is the regression curve given by Hyllienmark, Ludvigsson, Brismar (1995).

Supplementary Figure 3:

Panels A to D, scatter plots relating the HbA1C to the residual of the peroneal nerve conduction speed (dNCS). The colours in panels A to D reflect the antibody status of the patient with the relevant autoantibody or autoimmune disease given in the legend on the right of each panel.

Supplementary Figure 4:

Scatter plot relating the duration to the dNCS during the first five years, the dots are coloured according to the standard deviation of mean glucose. The blue line is the regression line (p = 0.0041).

Supplementary table 1: Raw data list of the diabetes group

Supplementary table 2: Statistics on NCS diabetes vs. control group

Supplementary table 3: Raw data list of the control group

Supplementary table 4: Summary of the control group

## Notes

### Competing Interest Statement

The authors have declared no competing interest.

### Funding Statement

This study did not receive any funding.

### Author Declarations

The study was approved by the ethics committee of Eastern Switzerland (Ethikkommission Ostschweiz, approval no: 22/018) and registered at the Swiss project database (2022-00216). Written informed consent was obtained from the caregivers before the inclusion of participants.

## REFERENCES

Barkai L, Kempler P. Puberty as a risk factor for diabetic neuropathy. Diabetes Care. 2000 Jul;23(7):1044–5.

Benjamini Y, Hochberg Y. Controlling the false discovery rate: a practical and powerful approach to multiple testing. J Roy Statistical Soc B. 1995;57:289–300.

Benbow SJ, Wallymahmed ME, MacFarlane IA. Diabetic peripheral neuropathy and quality of life. QJM. 1998 Nov;91(11):733–7.

Bierhaus A, Haslbeck KM, Humpert PM, Liliensiek B, Dehmer T, Morcos M et al. Loss of pain perception in diabetes is dependent on a receptor of the immunoglobulin superfamily. J Clin Invest. 2004 Dec;114(12):1741–51.

Bierhaus A, Schiekofer S, Schwaninger M, Andrassy M, Humpert PM, Chen J et al. Diabetes-associated sustained activation of the transcription factor nuclear factor-kappaB. Diabetes. 2001 Dec;50(12):2792–808.

Broser P, Luetschg J. Electromyography and electro-neurography in the neuropediatric diagnostic process. Klinische Neurophysiologie 2020;51:73–81.

Denys EH. AAEM minimonograph #14: The influence of temperature in clinical neurophysiology. Muscle Nerve. 1991 Sep;14(9):795–811.

DIAMOND Project Group. Incidence and trends of childhood Type 1 diabetes worldwide 1990-1999. Diabet Med. 2006 Aug;23(8):857–66.

Ekberg K, Brismar T, Johansson BL, Jonsson B, Lindström P, Wahren J. Amelioration of sensory nerve dysfunction by C-Peptide in patients with type 1 diabetes. Diabetes. 2003 Feb;52(2):536–41.

García-García A, Calleja-Fernández J. Neurophysiology of the development and maturation of the peripheral nervous system. Rev Neurol. 2004 Jan 1-15;38(1):79–83.

Gupta N, Arora M, Sharma R, Arora KS. Peripheral and Central Nervous System Involvement in Recently Diagnosed Cases of Hypothyroidism: An Electrophysiological Study. Ann Med Health Sci Res. 2016 Sep-Oct;6(5):261–266.

Hollander M and Wolfe DA. Nonparametric statistical methods. New York: John Wiley & Sons; 1973:27–33, 68–75.

Hyllienmark L, Ludvigsson J, Brismar T. Normal values of nerve conduction in children and adolescents. Electroencephalogr Clin Neurophysiol. 1995 Oct;97(5):208–14.

Hyllienmark L, Brismar T, Ludvigsson J. Subclinical nerve dysfunction in children and adolescents with IDDM. Diabetologia. 1995 Jun;38(6):685–92

Hyllienmark L, Alstrand N, Jonsson B, Ludvigsson J, Cooray G, Wahlberg-Topp J. Early electrophysiological abnormalities and clinical neuropathy: a prospective study in patients with type 1 diabetes. Diabetes Care. 2013 Oct;36(10):3187–94.

Johansson BL, Borg K, Fernqvist-Forbes E, Kernell A, Odergren T, Wahren J. Beneficial effects of C-peptide on incipient nephropathy and neuropathy in patients with Type 1 diabetes mellitus. Diabet Med. 2000 Mar;17(3):181–9.

Kallinikou D, Soldatou A, Tsentidis C, Louraki M, Kanaka-Gantenbein C, Kanavakis E, Karavanaki K. Diabetic neuropathy in children and adolescents with type 1 diabetes mellitus: Diagnosis, pathogenesis, and associated genetic markers. Diabetes Metab Res Rev. 2019 Oct;35(7):e3178.

Kamiya H, Zhang W, Sima AA. Dynamic changes of neuroskeletal proteins in DRGs underlie impaired axonal maturation and progressive axonal degeneration in type 1 diabetes. Exp Diabetes Res. 2009;2009:793281.

Kamiya H, Zhang W, Sima AA. The beneficial effects of C-Peptide on diabetic polyneuropathy. Rev Diabet Stud. 2009 Fall;6(3):187–202.

Klimontov VV, Saik OV, Korbut AI. Glucose Variability: How Does It Work? Int J Mol Sci. 2021 Jul 21;22(15):7783.

Ludvigsson J, Johannesson G, Heding L, Häger A, Larsson Y. Sensory nerve conduction velocity and vibratory sensibility in juvenile diabetics. Relationship to endogenous insulin. Acta Paediatr Scand. 1979 Sep;68(5):739–43.

Marcovecchio ML, Chiarelli F. Microvascular disease in children and adolescents with type 1 diabetes and obesity. Pediatr Nephrol. 2011 Mar;26(3):365–75.

Monti MC, Lonsdale JT, Montomoli C, Montross R, Schlag E, Greenberg DA. Familial risk factors for microvascular complications and differential male-female risk in a large cohort of American families with type 1 diabetes. J Clin Endocrinol Metab. 2007 Dec;92(12):4650–5.

Nathan DM, Genuth S, Lachin J, Cleary P, Crofford O, Davis M, Rand L, Siebert C, Diabetes Control and Complications Trial Research Group. The effect of intensive treatment of diabetes on the development and progression of long-term complications in insulin-dependent diabetes mellitus. N Engl J Med. 1993 Sep 30;329(14):977–86.

Olsen BS, Sjølie A, Hougaard P, Johannesen J, Borch-Johnsen K, Marinelli K et al. A 6-year nationwide cohort study of glycaemic control in young people with type 1 diabetes. Risk markers for the development of retinopathy, nephropathy and neuropathy. Danish Study Group of Diabetes in Childhood. J Diabetes Complications. 2000 Nov-Dec;14(6):295–300.

Patterson CC, Harjutsalo V, Rosenbauer J, Neu A, Cinek O, Skrivarhaug T et al. Trends and cyclical variation in the incidence of childhood type 1 diabetes in 26 European centres in the 25 year period 1989-2013: a multicentre prospective registration study. Diabetologia. 2019 Mar;62(3):408–417.

Pihoker C, Forsander G, Fantahun B, Virmani A, Corathers S, Benitez-Aguirre P, Fu J, Maahs DM. ISPAD Clinical Practice Consensus Guidelines 2018: The delivery of ambulatory diabetes care to children and adolescents with diabetes. Pediatr Diabetes. 2018 Oct;19 Suppl 27:84–104.

Solders G, Thalme B, Aguirre-Aquino M, Brandt L, Berg U, Persson A. Nerve conduction and autonomic nerve function in diabetic children. A 10-year follow-up study. Acta Paediatr. 1997 Apr;86(4):361–6.

Tinawi M. Disorders of Calcium Metabolism: Hypocalcemia and Hypercalcemia. Cureus. 2021 Jan 1;13(1):e12420.

R Core Team. R: a language and environment for statistical computing. Vienna, Austria. http://www.R-project.org/ (Accessed 23.07.2020)

Riihimaa PH, Suominen K, Tolonen U, Jäntti V, Knip M, Tapanainen P. Peripheral nerve function is increasingly impaired during puberty in adolescents with type 1 diabetes. Diabetes Care. 2001 Jun;24(6):1087–92.

Vague P, Coste TC, Jannot MF, Raccah D, Tsimaratos M. C-peptide, Na+,K(+)-ATPase, and diabetes. Exp Diabesity Res. 2004 Jan-Mar;5(1):37–50.

Verrotti A, Giuva PT, Morgese G, Chiarelli F. New trends in the etiopathogenesis of diabetic peripheral neuropathy. J Child Neurol. 2001 Jun;16(6):389–94.

Vlassara H, Brownlee M, Cerami A. Accumulation of diabetic rat peripheral nerve myelin by macrophages increases with the presence of advanced glycosylation endproducts. J Exp Med. 1984 Jul 1;160(1):197–207.

Washburn RL, Mueller K, Kaur G, Moreno T, Moustaid-Moussa N, Ramalingam L, Dufour JM. C-Peptide as a Therapy for Type 1 Diabetes Mellitus. Biomedicines. 2021 Mar 8;9(3):270.

White NH, Waltman SR, Krupin T, Santiago JV. Reversal of neuropathic and gastrointestinal complications related to diabetes mellitus in adolescents with improved metabolic control. J Pediatr. 1981 Jul;99(1):41–5.

Yagihashi S, Mizukami H, Sugimoto K. Mechanism of diabetic neuropathy: Where are we now and where to go? J Diabetes Investig. 2011 Jan 24;2(1):18–32.

Ziegler D, Behler M, Schroers-Teuber M, Roden M. Near-normoglycaemia and development of neuropathy: a 24-year prospective study from diagnosis of type 1 diabetes. BMJ Open. 2015 Jun 24;5(6):e006559.

