## Supplementary figures and images for "Peripheral nerve conduction speed shows a disease control-dependent and -independent drop in type 1 diabetes mellitus in children"

### Figure 1

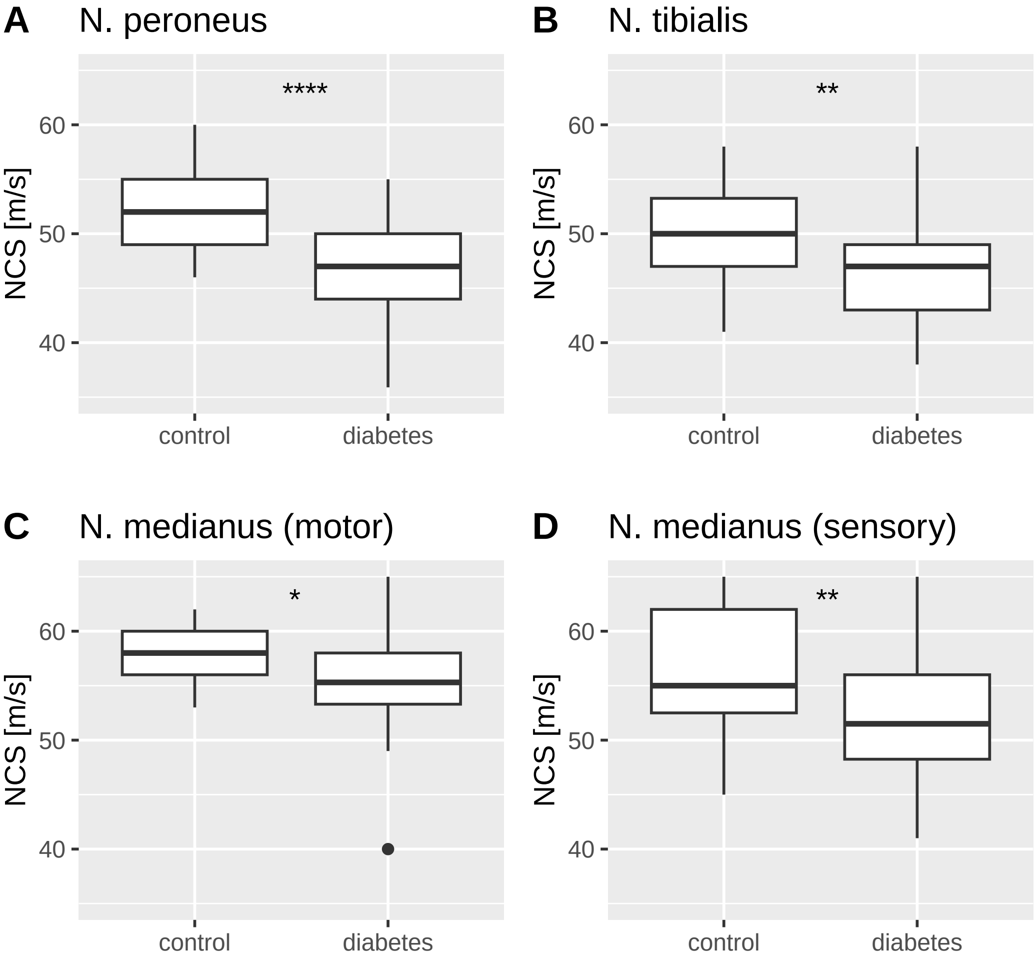

### Figure 2

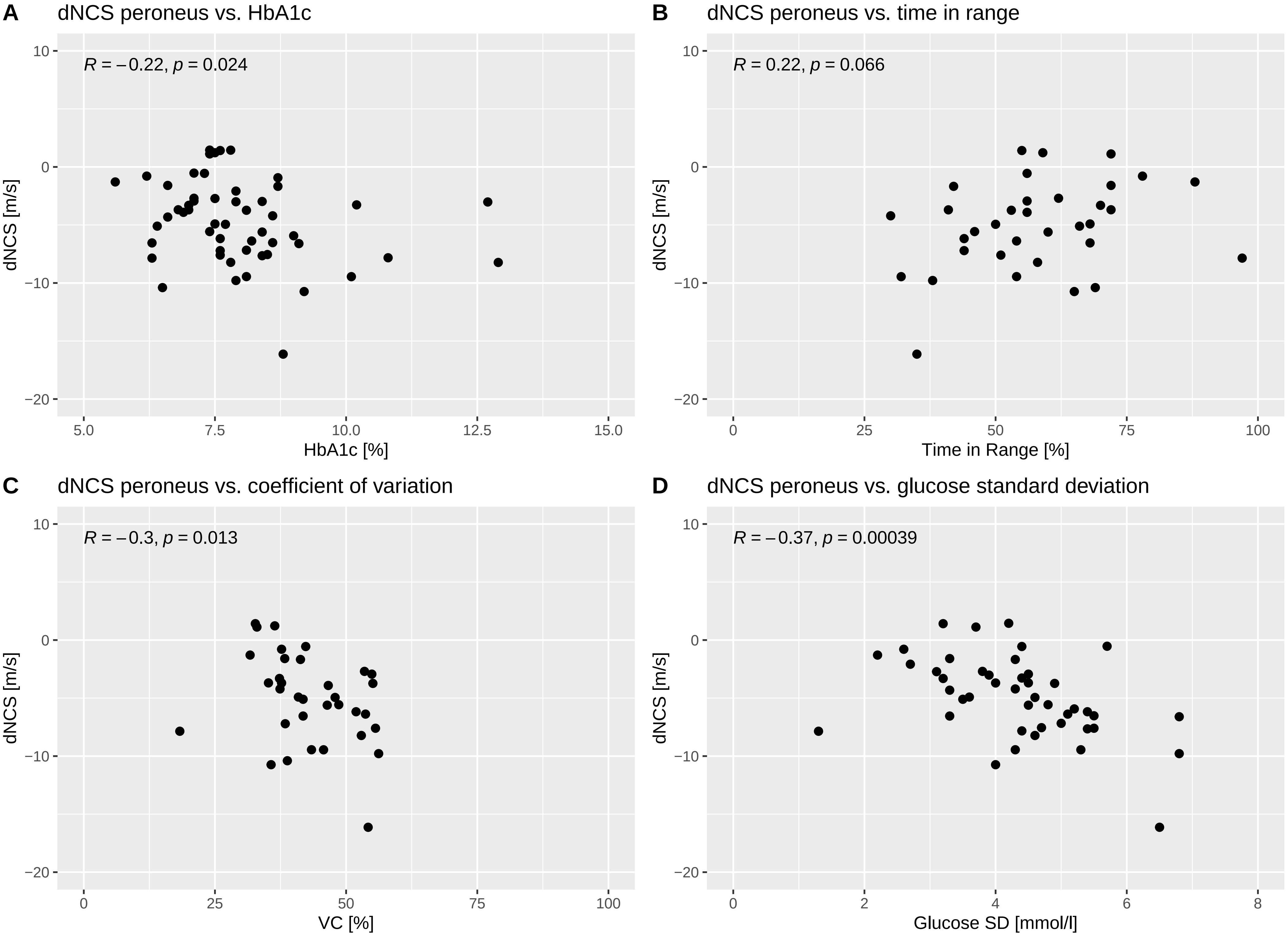

### Figure 3

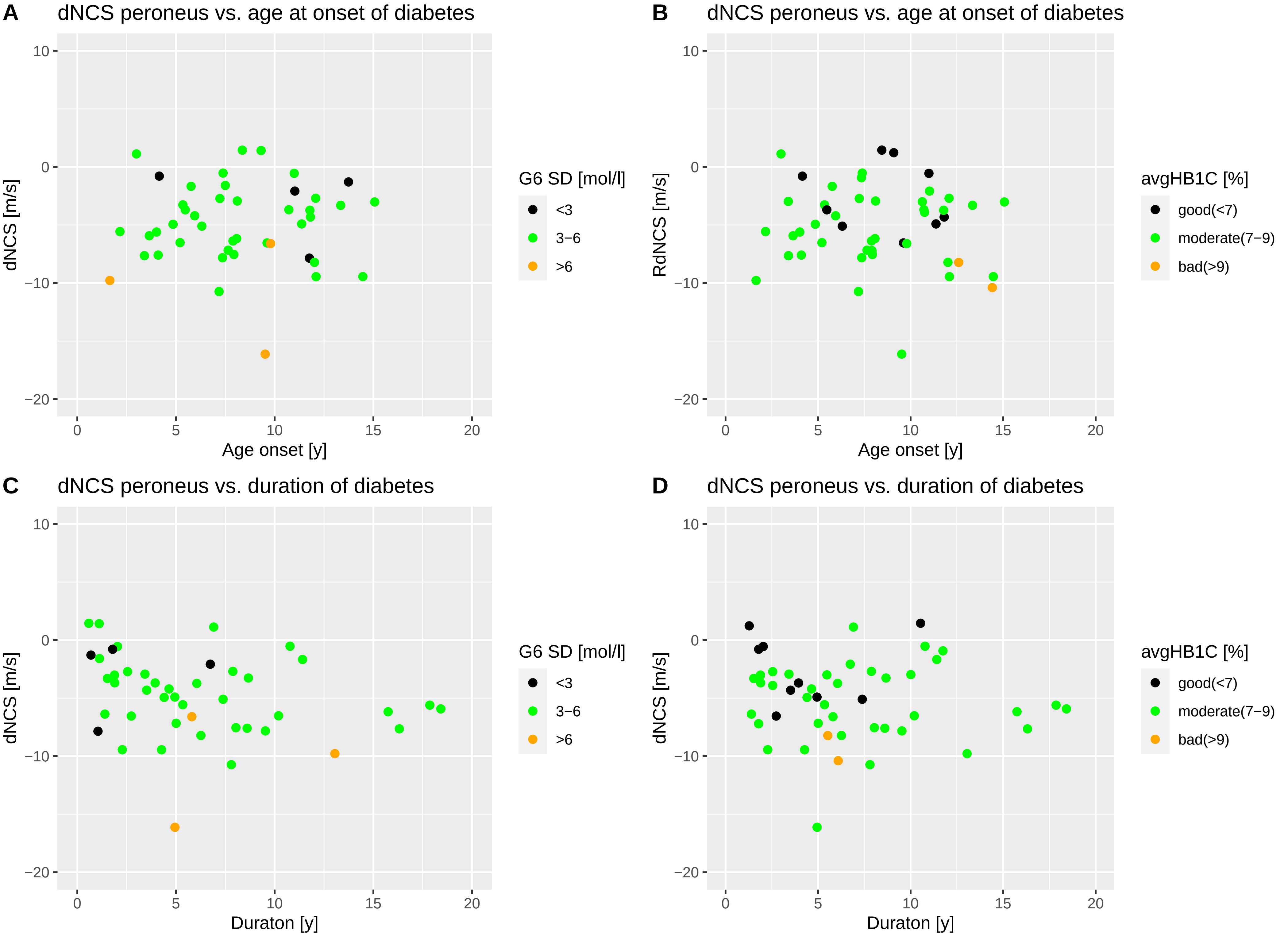

### Supplementary Figure 1

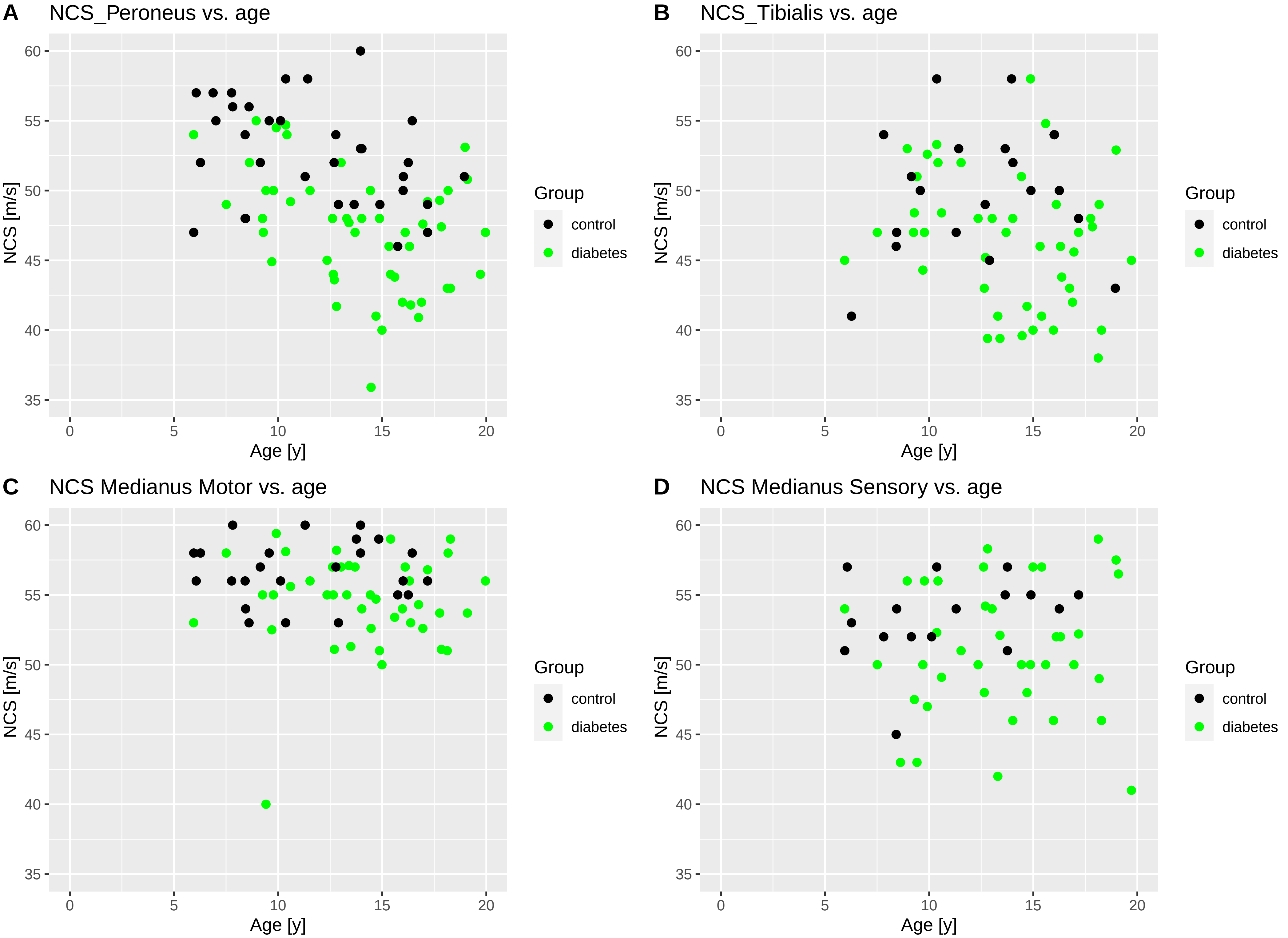

### Supplementary Figure 2

Supplementary Figure 2

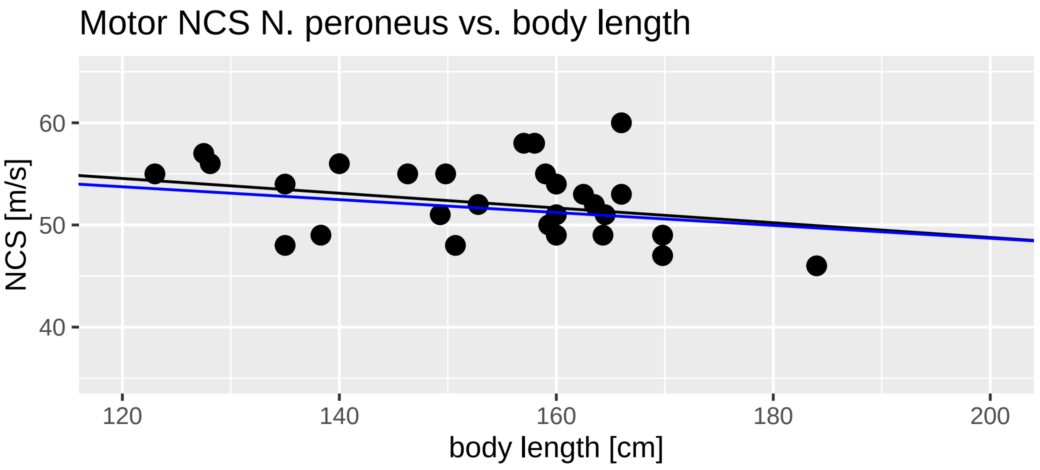

### Supplementary Figure 3

Supplementary Figure 3

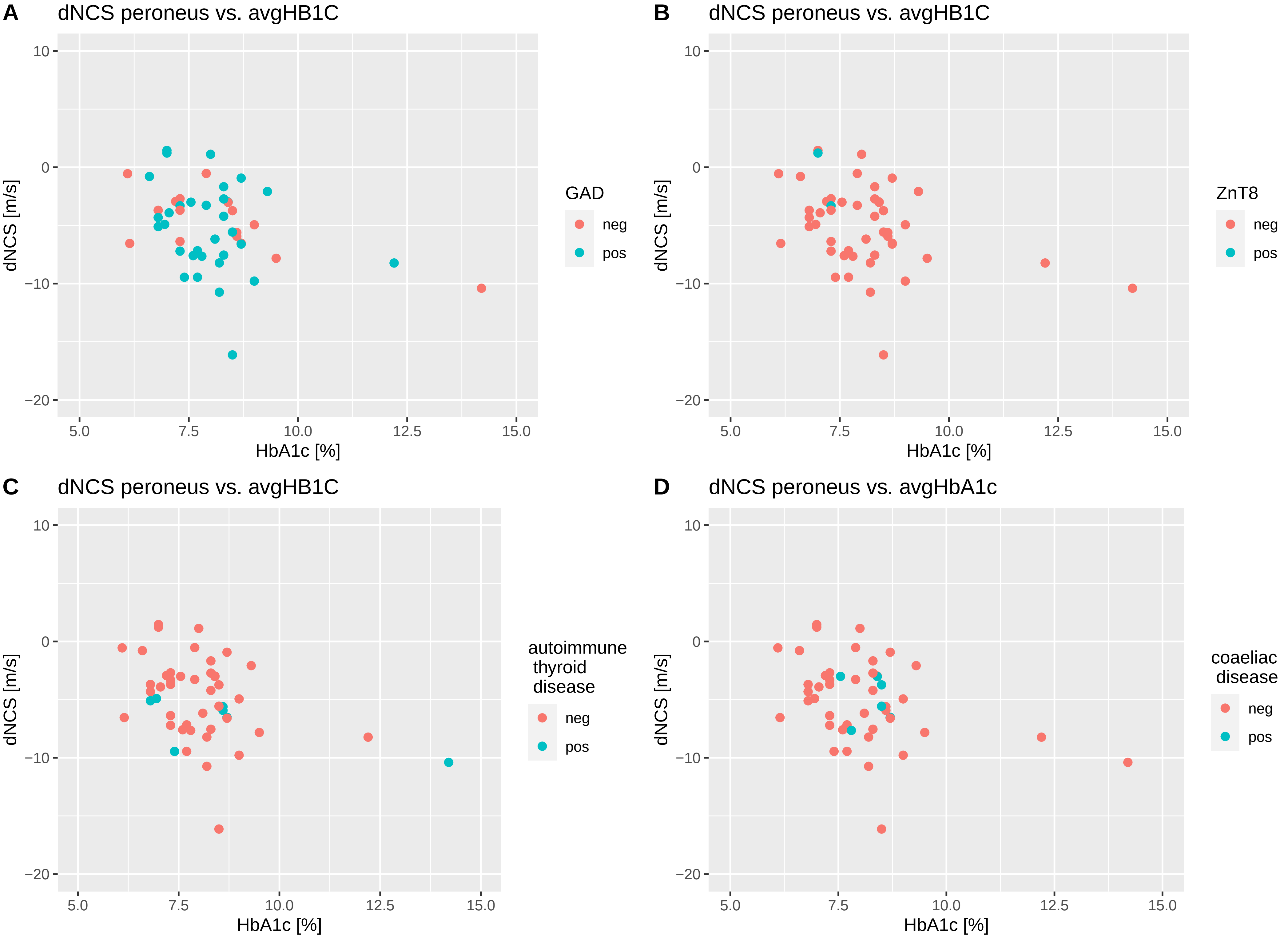

### Supplementary Figure 4

Supp. Figure 4: dNCS peroneus vs. duration of diabetes

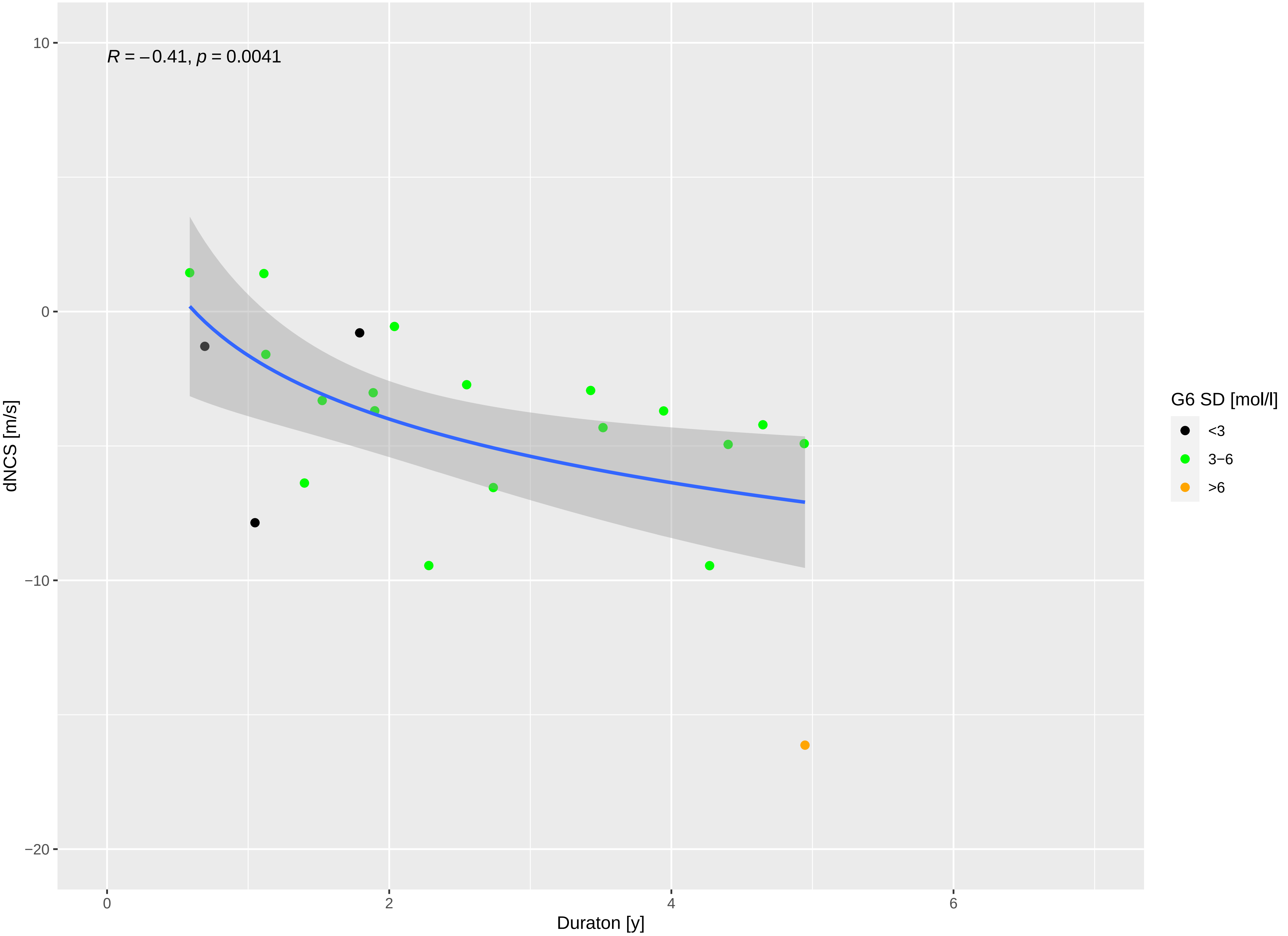
