## Supplementary material for "Peripheral nerve conduction speed shows a disease control-dependent and -independent drop in type 1 diabetes mellitus in children": Table 1

Table 1: Summary of Studydata (n=54, #Male=28, #PosAK=48, #Sensor=38, #Pump=28)

| Parameter | Min | Mean | Max |
|---|---|---|---|
| Age at onset (y) | 1.65 | 8.44 | 15.07 |
| Age at NCS (y) | 5.94 | 14.52 | 23.82 |
| Duration of DM (y) | 0.59 | 6.08 | 18.42 |
| Height (cm) | 120.40 | 160.90 | 186.50 |
| Weight (kg) | 20.70 | 57.02 | 94.30 |
| HbA1c (%) | 5.60 | 7.99 | 12.90 |
| Mean HbA1c last five years (%) | 6.10 | 8.04 | 14.20 |
| TIR (%) | 21.00 | 54.78 | 97.00 |
| Mean glucose (mmol/l) | 4.50 | 9.75 | 15.70 |
| SD of mean glucose (mmol/l) | 1.30 | 4.37 | 6.80 |
| Coefficient of variation (%) | 18.30 | 43.60 | 69.60 |
