## Supplementary Table 1 for "Peripheral nerve conduction speed shows a disease control-dependent and -independent drop in type 1 diabetes mellitus in children"

Supplementary Table 1: Raw Data

| Demographics | | | Diabetes Data | | | | NCS Data | | |
|---|---|---|---|---|---|---|---|---|---|
| ID | age onset [y] | age NCS [y] | duration DBM [y] | HB1c at NCS [%] | mean 5y HB1c [%] | betacell antibodies | peroneus [m/s] | tibial [m/s] | median [m/s] |
| 28 | 0-5 | 5-10 | 1.8 | 6.20 | 6.60 | pos | 54.0 | 45.0 | 53.0 |
| 24 | 0-5 | 5-10 | 5.3 | 7.40 | 8.50 | pos | 49.0 | 47.0 | 58.0 |
| 5 | 5-10 | 5-10 | 1.1 | 6.60 | — | pos | 52.0 | — | 61.0 |
| 46 | 5-10 | 5-10 | 0.6 | 7.80 | — | pos | 55.0 | 53.0 | 62.0 |
| 57 | 0-5 | 5-10 | 4.4 | 7.70 | 9.00 | pos | 48.0 | 47.0 | 55.0 |
| 10 | 5-10 | 5-10 | 1.4 | 8.20 | 7.30 | pos | 47.0 | 48.4 | 60.9 |
| 9 | 5-10 | 5-10 | 3.9 | 7.00 | 6.80 | neg | 50.0 | 51.0 | 40.0 |
| 34 | 5-10 | 5-10 | 1.8 | 7.60 | 7.30 | pos | 44.9 | 44.3 | 52.5 |
| 42 | 5-10 | 5-10 | 2.5 | 7.50 | 8.30 | pos | 50.0 | 47.0 | 55.0 |
| 52 | 0-5 | 5-10 | 6.9 | 7.40 | 8.00 | pos | 54.5 | 52.6 | 59.4 |
| 44 | 5-10 | 10-15 | 1.3 | 7.50 | 7.00 | pos | 54.7 | 53.3 | 58.1 |
| 26 | 5-10 | 10-15 | 1.1 | 7.60 | — | pos | 54.0 | 52.0 | 63.0 |
| 51 | 5-10 | 10-15 | 4.6 | 8.60 | 8.30 | pos | 49.2 | 48.4 | 55.6 |
| 50 | 5-10 | 10-15 | 3.4 | 7.10 | 7.20 | neg | 50.0 | 52.0 | 56.0 |
| 48 | 5-10 | 10-15 | 2.7 | 6.30 | 6.15 | pos | 45.0 | 48.0 | 55.0 |
| 22 | 10-15 | 10-15 | 1.9 | 6.80 | 7.30 | pos | 48.0 | — | 57.0 |
| 41 | 5-10 | 10-15 | 5.0 | 8.10 | 7.70 | pos | 44.0 | 43.0 | 55.0 |
| 32 | 0-5 | 10-15 | 8.6 | 7.60 | 7.60 | pos | 43.6 | 45.2 | 51.1 |

Supplementary Table 1: Raw Data

| | | | | | | | | | |
|---|---|---|---|---|---|---|---|---|---|
| 14 | 10-15 | 10-15 | 1.0 | 6.30 | — | pos | 41.7 | 39.4 | 58.2 |
| 49 | 10-15 | 10-15 | 2.0 | 7.30 | 6.10 | neg | 52.0 | 48.0 | 57.0 |
| 25 | 10-15 | 10-15 | 2.5 | 6.90 | 7.05 | pos | 48.0 | 41.0 | 55.0 |
| 15 | 0-5 | 10-15 | 10.0 | 8.40 | 8.40 | neg | 47.7 | 39.4 | 57.1 |
| 33 | 10-15 | 10-15 | 2.6 | 9.20 | 6.95 | pos | — | — | 51.3 |
| 45 | 5-10 | 10-15 | 7.4 | 6.40 | 6.80 | pos | 47.0 | 47.0 | 57.0 |
| 8 | 5-10 | 10-15 | 8.7 | 10.20 | 7.90 | pos | 48.0 | 48.0 | 54.0 |
| 55 | 10-15 | 10-15 | 0.7 | 5.60 | — | pos | 50.0 | 51.0 | 55.0 |
| 43 | 5-10 | 10-15 | 4.9 | 8.80 | 8.50 | pos | 35.9 | 39.6 | 52.6 |
| 6 | 0-5 | 10-15 | 13.1 | 7.90 | 9.00 | pos | 41.0 | 41.7 | 54.7 |
| 11 | 10-15 | 10-15 | 1.5 | 7.00 | 7.30 | pos | 48.0 | 58.0 | 51.0 |
| 21 | 5-10 | 10-15 | 7.8 | 9.20 | 8.20 | pos | 40.0 | 40.0 | 50.0 |
| 29 | 10-15 | 15-20 | 3.5 | 6.60 | 6.80 | pos | 46.0 | 46.0 | 68.0 |
| 56 | 5-10 | 15-20 | 10.2 | 8.60 | 8.70 | neg | 44.0 | 41.0 | 59.0 |
| 39 | 5-10 | 15-20 | 5.8 | 9.10 | 8.70 | pos | 43.8 | 54.8 | 53.4 |
| 12 | 5-10 | 15-20 | 8.0 | 8.50 | 8.30 | pos | 42.0 | 40.0 | 54.0 |
| 27 | 10-15 | 15-20 | 5.5 | 7.90 | 7.55 | pos | 47.0 | 49.0 | 57.0 |

Supplementary Table 1: Raw Data

| | | | | | | | | | |
|---|---|---|---|---|---|---|---|---|---|
| 3 | 10-15 | 15-20 | 4.9 | 7.50 | 6.95 | pos | 46.0 | 46.0 | 56.0 |
| 36 | 10-15 | 15-20 | 4.3 | 10.10 | 7.40 | pos | 41.8 | 43.8 | 53.0 |
| 35 | 10-15 | 15-20 | 2.3 | 8.10 | 7.70 | pos | 40.9 | 43.0 | 54.3 |
| 1 | 5-10 | 15-20 | 9.5 | 10.80 | 9.50 | pos | 42.0 | 42.0 | NA |
| 19 | 15-20 | 15-20 | 1.9 | 12.70 | 8.40 | pos | 47.6 | 45.6 | 52.6 |
| 16 | 5-10 | 15-20 | 11.4 | 8.70 | 8.30 | pos | 49.2 | 47.0 | 56.8 |
| 37 | 10-15 | 15-20 | 6.7 | 7.90 | 9.30 | pos | 49.3 | 48.0 | 53.7 |
| 40 | 10-15 | 15-20 | 6.1 | 8.10 | 8.50 | pos | 47.4 | 47.4 | 51.1 |
| 58 | 10-15 | 15-20 | 5.5 | 12.90 | 12.20 | pos | 43.0 | 38.0 | 51.0 |
| 13 | 5-10 | 15-20 | 10.8 | 7.10 | 7.90 | pos | 50.0 | 49.0 | 58.0 |
| 18 | 10-15 | 15-20 | 6.3 | 7.80 | 8.20 | pos | 43.0 | 40.0 | 59.0 |
| 30 | 5-10 | 15-20 | 10.5 | 7.40 | 7.00 | pos | 53.1 | 52.9 | 62.2 |
| 38 | 5-10 | 15-20 | 11.7 | 8.70 | 8.70 | pos | 50.8 | — | 53.7 |
| 54 | 0-5 | 15-20 | 16.3 | 8.40 | 7.80 | pos | 44.0 | 45.0 | 65.0 |
| 4 | 10-15 | 15-20 | 7.9 | 7.10 | 7.30 | pos | 47.0 | — | 56.0 |
| 7 | 10-15 | 20-25 | 6.1 | 6.50 | 14.20 | pos | 40.0 | 49.0 | 49.0 |
| 17 | 0-5 | 20-25 | 17.9 | 8.40 | 8.60 | pos | 44.2 | — | 56.1 |

Supplementary Table 1: Raw Data

| | | | | | | | | | | |
|---|---|---|---|---|---|---|---|---|---|---|
| 2 | 0-5 | 20-25 | 18.4 | 9.00 | 8.60 | neg | | 45.6 | 45.9 | 59.0 |
| 31 | 5-10 | 20-25 | 15.7 | 7.60 | 8.10 | pos | | 44.0 | 45.0 | 58.0 |
