## Supplementary Table 2 for "Peripheral nerve conduction speed shows a disease control-dependent and -independent drop in type 1 diabetes mellitus in children"

Supplementary Table 2: Statistic NCS Diabetes vs. Control

| Nerve | diabetes gr. [m/s] | control gr. [m/s] | p-Value | | n Control | n Diabetes |
|---|---|---|---|---|---|---|
| N.peroneus | 47.0 | 52 | 0.0000 | **** | 33 | 53 |
| N.tibialis | 47.0 | 50 | 0.0042 | ** | 20 | 48 |
| N.medianus (m) | 55.6 | 58 | 0.0281 | * | 29 | 53 |
| N.medianus (s) | 51.5 | 55 | 0.0019 | ** | 24 | 46 |
