## Supplementary Table 3 for "Peripheral nerve conduction speed shows a disease control-dependent and -independent drop in type 1 diabetes mellitus in children"

Supp. Table 3:Control Data Study Group

| | | | | | right | | | | |
|---|---|---|---|---|---|---|---|---|---|
| **Index** | **Age [y]** | **Sex** | **Height [cm]** | **Weight [kg]** | **N.peroneus [m/s]** | **N.tibialis [m/s]** | **N.medianus (m) [m/s]** | **N.medianus (s) [m/s]** | **N.pero** |
| 33 | 5-10 | m | 111.00 | 16.20 | 47.00 | | | | |
| 28 | 5-10 | w | 116.70 | 21.50 | 57.00 | | | | |
| 11 | 5-10 | m | 111.00 | 16.70 | 52.00 | 41.00 | 58.00 | 53.00 | |
| 1 | 5-10 | w | 119.00 | 22.70 | | | | | |
| 22 | 5-10 | m | 123.00 | 22.00 | 55.00 | | | | |
| 26 | 5-10 | w | 127.50 | 33.40 | 57.00 | | | | |
| 4 | 5-10 | w | 140.00 | | 56.00 | 54.00 | 60.00 | 52.00 | |
| 32 | 5-10 | m | 135.00 | 28.00 | 48.00 | 46.00 | | | |
| 3 | 5-10 | m | 150.70 | 44.50 | | | | | |
| 27 | 5-10 | w | 128.10 | 25.60 | 56.00 | | 53.00 | 64.00 | |
| 31 | 5-10 | m | 152.80 | 48.80 | 52.00 | 51.00 | 57.00 | 52.00 | |
| 20 | 5-10 | w | 146.30 | 55.00 | 55.00 | 50.00 | 58.00 | 64.00 | |
| 24 | 10-15 | w | 149.80 | 36.40 | 55.00 | | 56.00 | 52.00 | |
| 35 | 10-15 | m | 158.00 | 44.00 | 58.00 | 58.00 | 53.00 | 57.00 | |
| 30 | 10-15 | w | 149.30 | 40.00 | 51.00 | 47.00 | 60.00 | 54.00 | |

Supp. Table 3:Control Data Study Group

| | | | | | | | | | |
|---|---|---|---|---|---|---|---|---|---|
| 21 | 10-15 | w | 157.00 | 35.50 | 58.00 | 53.00 | 61.00 | 63.00 | |
| 10 | 10-15 | w | | 9.00 | | | | | |
| 23 | 10-15 | w | 160.00 | 64.30 | 54.00 | | 57.00 | | |
| 17 | 10-15 | w | 138.30 | 32.80 | 49.00 | 45.00 | 53.00 | 61.00 | |
| 7 | 10-15 | m | 160.00 | 53.00 | 49.00 | 53.00 | 61.00 | 55.00 | |
| 5 | 10-15 | w | 161.00 | 62.00 | | | 59.00 | 51.00 | |
| 18 | 10-15 | w | 166.00 | 61.30 | 53.00 | 58.00 | 60.00 | 63.00 | [illegible] |
| 29 | 10-15 | w | 162.50 | 69.50 | | | | | |
| 12 | 10-15 | w | 155.00 | 38.00 | | | 59.00 | | |
| 15 | 10-15 | w | 164.30 | 56.00 | 49.00 | 50.00 | 62.00 | 55.00 | |
| 13 | 15-20 | m | 184.00 | 65.90 | | | 55.00 | | [illegible] |
| 19 | 15-20 | w | 159.30 | 42.40 | 50.00 | 54.00 | 56.00 | 63.00 | |
| 2 | 15-20 | w | 164.50 | 51.00 | 51.00 | 54.00 | | | |
| 16 | 15-20 | m | 163.50 | 59.30 | 52.00 | 50.00 | | | |
| 34 | 15-20 | m | 159.00 | 55.00 | | | | | |
| 25 | 15-20 | w | 169.80 | 68.70 | 47.00 | 48.00 | 56.00 | 55.00 | [illegible] |
| 6 | 15-20 | w | 160.00 | 55.00 | 51.00 | 43.00 | | | |
