## Supplementary Table 4 for "Peripheral nerve conduction speed shows a disease control-dependent and -independent drop in type 1 diabetes mellitus in children"

Supp. Table 4: Summary of control data (n=32, #Male=11)

| Parameter | Min | Mean | Max |
|---|---|---|---|
| Age at NCS (y) | 5.95 | 11.73 | 18.95 |
| Height at NCS (cm) | 111.00 | 148.46 | 184.00 |
| Weight at NCS (cm) | 9.00 | 43.02 | 69.50 |
